# Heart injury signs are associated with higher and earlier mortality in coronavirus disease 2019 (COVID-19)

**DOI:** 10.1101/2020.02.26.20028589

**Authors:** Chaomin Wu, Xianglin Hu, Jianxin Song, Chunling Du, Jie Xu, Dong Yang, Dechang Chen, Ming Zhong, Jinjun Jiang, Weining Xiong, Ke Lang, Yuye Zhang, Guohua Shi, Lei Xu, Yuanlin Song, Xin Zhou, Ming Wei, Junhua Zheng, The first batch of medical teams from Shanghai to support Hubei, China and study group

## Abstract

**Importance:** Heart injury can be easily induced by viral infection such as adenovirus and enterovirus. However, whether coronavirus disease 2019 (COVID-19) causes heart injury and hereby impacts mortality has not yet been fully evaluated.

**Objective:** To explore whether heart injury occurs in COVID-19 on admission and hereby aggravates mortality later.

**Design, Setting, and Participants:** A single-center retrospective cohort study including 188 COVID-19 patients admitted from December 25, 2019 to January 27, 2020 in Wuhan Jinyintan Hospital, China; follow up was completed on February 11, 2020.

**Exposures:** High levels of heart injury indicators on admission (hs-TNI; CK; CK-MB; LDH; α-HBDH).

**Main Outcomes and Measures:** Mortality in hospital and days from admission to mortality (survival days).

**Results:** Of 188 patients with COVID-19, the mean age was 51.9 years (standard deviation: 14.26; range: 21∼83 years) and 119 (63.3%) were male. Increased hs-TnI levels on admission tended to occur in older patients and patients with comorbidity (especially hypertension). High hs-TnI on admission (≥ 6.126 pg/mL), even within the clinical normal range (0∼28 pg/mL), already can be associated with higher mortality. High hs-TnI was associated with increased inflammatory levels (neutrophils, IL-6, CRP, and PCT) and decreased immune levels (lymphocytes, monocytes, and CD4^+^ and CD8^+^ T cells). CK was not associated with mortality. Increased CK-MB levels tended to occur in male patients and patients with current smoking. High CK-MB on admission was associated with higher mortality. High CK-MB was associated with increased inflammatory levels and decreased lymphocytes. Increased LDH and α-HBDH levels tended to occur in older patients and patients with hypertension. Both high LDH and α-HBDH on admission were associated with higher mortality. Both high LDH and α-HBDH were associated with increased inflammatory levels and decreased immune levels. hs-TNI level on admission was negatively correlated with survival days (*r*= -0.42, 95% CI= -0.64∼-0.12, P=0.005). LDH level on admission was negatively correlated with survival days (*r*= -0.35, 95% CI= -0.59∼-0.05, P=0.022).

**Conclusions and Relevance:** Heart injury signs arise in COVID-19, especially in older patients, patients with hypertension and male patients with current smoking. COVID-19 virus might attack heart via inducing inflammatory storm. High levels of heart injury indicators on admission are associated with higher mortality and shorter survival days. COVID-19 patients with signs of heart injury on admission must be early identified and carefully managed by cardiologists, because COVID-19 is never just confined to respiratory injury.

**Key points:** *Question:* Does coronavirus disease 2019 (COVID-19) cause heart injury and hereby impact mortality?

*Findings:* In this retrospective cohort study including 188 patients with COVID-19, patients with high levels of high-sensitivity cardiac troponin I (hs-TNI) on admission had significantly higher mortality (50.0%) than patients with moderate or low levels of hs-TNI (10.0% or 9.1%). hs-TNI level on admission was significantly negatively correlated with survival days (*r*= -0.42, 95% CI= -0.64∼-0.12, P=0.005).

*Meaning:* COVID-19 patients with signs of heart injury on admission must be early identified and carefully managed by cardiologists, in order to maximally prevent or rescue heart injury-related mortality in COVID-19.

## Introduction

Viral infection is a common cause of heart injury such as myocarditis. It is well known that adenovirus, enterovirus and herpesvirus are the most common infectious etiologies for heart injury [1]. Since coronavirus-related carditis was first reported in 1980 [2], increasing studies noted that coronavirus is an uncommon but unneglectable pathogen for heart injury. For instance, we found the coronavirus of Severe Acute Respiratory Syndrome Coronavirus (SARS-CoV) in 2003 in China had no obvious connections with heart injury. However, it was reported that the coronavirus of Middle East Respiratory Syndrome (MERS-CoV) in 2012 in Saudi Arabia could cause acute myocarditis and heart failure [3].

Coronavirus disease 2019 (COVID-19) is a novel infectious viral pneumonia breaking out in Wuhan, China since Dec, 2019 [4, 5]. By Feb 20, 2020 in China, 75,465 patients were diagnosed with COVID-19 and 2,236 patients deceased [6]. Based on our recent experts clinical experience on COVID-19, we found severe COVID-19 patients are difficult to rescue not only because respiratory failure but also susceptibility to heart failure. The coronavirus for COVID-19 is a new strain which was not previously identified. Whether the coronavirus for COVID-19 causes heart injury and thereby impacts mortality has not been fully evaluated. Therefore, in this retrospective cohort study, we fully assessed heart injury indicators on admission in COVID-19 patients. We aimed to explore whether heart injury already occurs in COVID-19 at early stage and hereby aggravates mortality later.

## Methods

### Study population

From Dec 25, 2019 to Jan 27, 2020, 188 COVID-19 patients admitted to Wuhan Jinyintan Hospital (Hubei, China) were selected. All enrolled patients were definitely diagnosed with COVID-19 pneumonia according to the previously reported criteria [7]. Patients with missing clinical information were excluded. Age of the enrolled patients ranged from 21 to 83 years old. Up to Feb 11, 2020, follow up in hospital was completed for all patients. One hundred and forty-five patients were cured and discharged while 43 patients deceased in hospital. This research was approved by the Review Board of Wuhan Jinyintan Hospital. Written informed consent was waived because this research was retrospective observation and deindividuation.

### Data collection

Data were retrieved from electronic medical records. Demographic and clinical parameters included age, gender, history of Huanan seafood market exposure, history of family infection, smoking status, comorbidity, initial symptoms and signs, vital signs on admission, complications on admission and drug treatment regimen. Primary outcome was mortality in hospital. Days from admission to death (survival days) were also calculated for deceased patients. Whether transfer to intensive care unit (ICU) was recorded as a secondary outcome.

Heart injury indicators measured on admission were collected, including high-sensitivity cardiac troponin I (hs-TNI), creatine kinase (CK), creatine kinase-MB (CK-MB), lactic dehydrogenase (LDH) and α-hydroxybutyrate dehydrogenase (α-HBDH). The clinical normal range of hs-TNI was 0∼28 pg/mL. The clinical normal range of CK was 50∼310 U/L. The clinical normal range of CK-MB was 0∼24 U/L. The clinical normal range of LDH was 120∼250 U/L. The clinical normal range of α-HBDH was 72∼182 U/L. All Heart injury indicators were divided into three levels (low, moderate or high level) based on the cutoff values of 33^th^ and 67^th^ percentile.

Inflammatory levels indicators were collected, including white blood cells (WBC), neutrophils, interleukin-6 (IL-6), C-reaction protein (CRP), procalcitonin (PCT) and erythrocyte sedimentation rate (ESR). Immune levels indicators were collected, including lymphocytes, monocytes, T cells count, and CD4^+^ and CD8^+^ T cells count.

### Statistical analysis

SPSS for Windows (Version 24.0, IBM) and Graphpad prism 7.0 software were used for statistical analysis. Continuous parameters were displayed by mean ± standard deviation (SD) or median with quartiles. Categorical parameters were displayed by percentage with number. One way ANOVA or Kruskal-Wallis test was performed for multiple comparisons of continuous data. Tukey’s multiple comparisons test or Dunn’s multiple comparisons test was used for adjusted P value. Chi-square test was performed for categorical data and Bonferroni method was used for adjusted P value. Spearman correlation was performed. Coefficient of *r* with 95% confidence interval (CI) was used to measure the correlation magnitude. P< 0.05 was considered statistically significant.

## Results

The baseline clinical and laboratory characteristics were presented in **Table 1**. Patients’ mean age was 51.9 ± 14.26 years old. Male accounted for 63.3% of the population. Huanan seafood market exposure accounted for 48.9% while infection by family members accounted for 8.0% of the population. There were 9.0% patients with current smoking. Hypertension (20.2%) and diabetes (10.6%) ranked as the most common comorbidities. Fever (92.6%) and cough (83.5%) were the most common initial symptoms and signs. Approximately 11.2% patients had hs-TNI exceeding clinical upper normal limit on admission. Approximately 68.6% patients had LDH exceeding clinical upper normal limit on admission. Approximately 76.1% patients had α-HBDH exceeding clinical upper normal limit on admission. Approximately 11.2% patients had CK exceeding clinical upper normal limit on admission. Approximately 10.1% patients had CK-MB exceeding clinical upper normal limit on admission. Antibiotics (98.4%) and non-specific antiviral drugs (84.0%) were used in most patients. There were 50 (26.6%) patients transferring to ICU. There were 43 (22.9%) patients deceasing in hospital. The median survival days for deceased patients were 7 (4-11) days.

**Table 1.** Clinical and laboratory characteristics of the enrolled patients (n=188).

| Parameters |  | All patients |
| --- | --- | --- |
| Age (years) | Mean ± SD | 51.9± 14.26 |
|  | Median (25<sup>th</sup> and 75<sup>th</sup> percentile) | 51 (43~60) |
| Male |  | 119 (63.3%) |
| Huanan seafood market exposure |  | 92 (48.9%) |
| Infection by family members |  | 15 (8.0%) |
| Current smoking |  | 17 (9.0%) |
| Comorbidity <sup>a</sup> |  | 64 (34.0%) |
|  | Hypertension | 38 (20.2%) |
|  | Diabetes | 20 (10.6%) |
| Initial symptoms and signs <sup>b</sup> |  |  |
|  | Fever | 174 (92.6%) |
| | Highest temperature (°C) (n=156) | 38.73± 0.67 |
|  | Cough | 157 (83.5%) |
|  | Sputum | 74 (39.4%) |
|  | Breathlessness | 72 (38.3%) |
|  | Chest distress or pain | 63 (33.5%) |
|  | Fatigue or myalgia | 61 (32.5%) |
|  | Dyspnea | 24 (12.8%) |
| Days from symptoms/signs to admission |  | 9 (7-12) |
| Vital signs on admission |  |  |
| | Temperature (°C) | 36.91 ± 0.76 |
|  | Heart rate (beats/min) | 88 (82~98) |
|  | Respiratory rate (breaths/min) | 20 (20~23.75) |
|  | Systolic pressure (mmHg) | 122 (115~134.8) |
|  | Diastolic pressure (mmHg) | 75 (75~85) |
| Complications on admission |  |  |
|  | Respiratory failure | 20 (10.6%) |
|  | ARDS or sepsis | 6 (3.2%) |
| hs-TnI (pg/mL) | Median (25<sup>th</sup> and 75<sup>th</sup> percentile) | 3.65 (1.3~9.425) |
|  | 33<sup>th</sup> and 67<sup>th</sup> percentile | 2~6.126 |
|  | > 28 pg/mL ( clinical upper normal limit) | 21 (11.2%) |
| LDH (U/L) | Median (25<sup>th</sup> and 75<sup>th</sup> percentile) | 308.5 (233.3~389.8) |
|  | 33<sup>th</sup> and 67<sup>th</sup> percentile | 253.2~354.5 |
|  | > 250 U/L (clinical upper normal limit) | 129 (68.6%) |
| α-HBDH (n=173)<br>(U/L) | Median (25<sup>th</sup> and 75<sup>th</sup> percentile) | 255 (198.5~337) |
|  | 33<sup>th</sup> and 67<sup>th</sup> percentile | 215.8~305 |
|  | > 182 U/L (clinical upper normal limit) | 143 (76.1%) |
| CK (U/L) | Median (25<sup>th</sup> and 75<sup>th</sup> percentile) | 83 (46.25~159.5) |
|  | 33<sup>th</sup> and 67<sup>th</sup> percentile | 53.74~123.6 |
|  | > 310 U/L (clinical upper normal limit) | 21 (11.2%) |
| CK-MB (U/L) | Median (25<sup>th</sup> and 75<sup>th</sup> percentile) | 15 (12~20) |
|  | 33<sup>th</sup> and 67<sup>th</sup> percentile | 13~18 |
|  | > 24 U/L ( clinical upper normal limit) | 19 (10.1%) |
| Drug therapy |  |  |
|  | Antibiotics | 185 (98.4%) |
|  | Antiviral drugs | 158 (84.0%) |
|  | Corticosteroids | 59 (31.4%) |
| Transfer to ICU |  | 50 (26.6%) |
| Cured or deceased |  |  |
|  | Cured and discharged | 145 (77.1%) |
|  | Length of stay (days) for cured patients | 12 (9-14) |
|  | Deceased in hospital | 43 (22.9%) |
|  | Length of stay (days) for deceased patients<br>(survival days) | 7 (4-11) |

<sup>a</sup> Besides hypertension and diabetes, other comorbidity included chronic liver disease (9 cases), coronary heart disease (6 cases), cerebrovascular disease (6 cases), Parkinson's disease (3 cases), rheumatoid disease (3 cases), chronic kidney disease (3 cases), thyroid disease (2 cases), stomach disease (3 cases), chronic obstructive pulmonary disease (2 cases) and pulmonary tuberculosis (3 cases).
<sup>b</sup> Other symptoms and signs included headache (12 cases), nasal obstruction or nasal discharge (8 cases), nausea and vomiting (7 cases), dizziness (6 cases), pharyngalgia (6 cases) and diarrhea (2 cases).
Continuous data were presented by mean ± standard deviation (SD), median (25<sup>th</sup> to 75<sup>th</sup> percentile) or 33<sup>th</sup> to 67<sup>th</sup> percentile.
Categorical data were presented by number with percentage.
Abbreviations: ARDS: acute respiratory distress syndrome; hsTnI: High-sensitivity troponin I; LDH: Lactic dehydrogenase; α-HBDH: α-hydroxybutyrate dehydrogenase; CK: Creatine kinase; CK-MB: creatine kinase-MB; ICU: intensive care unit.

As shown in **Table 2**, increased hs-TnI levels were more likely to occur in older patients and patients with hypertension or diabetes. High hs-TnI was independent of acute respiratory symptoms and respiratory failure (*P*> 0.05). High hs-TnI on admission (≥ 6.126 pg/mL), even within the clinical normal range (0∼28 pg/mL), already can be associated with higher mortality (50.0%). High hs-TnI was significantly associated with increased inflammatory levels (neutrophils, IL-6, CRP, and PCT) and decreased immune levels (lymphocytes, monocytes, and CD4^+^ and CD8^+^ T cells). Survival days were obviously shorter for deceased patients with high hs-TNI (median: 7 days) when compared to patients with low and moderate hs-TNI (median: 11 and 8.5 days, respectively), although it was hard to reach statistical significance because the deceased cases in the three groups were extremely uneven distribution.

**Table 2.** Clinical characteristics and outcomes between patients with different hs-TnI levels.

| Parameters | Low hs-TnI group | Moderate hs-TnI group | High hs-TnI group |
| --- | --- | --- | --- |
| Age (years) | 43.57 ± 11.99 | 50.76 ± 11.11 <sup>#</sup> | 61.19 ± 13.98 * <sup>§</sup> |
| Male | 56.7% (34/60) | 69.7% (46/66) | 62.9% (39/62) |
| Current smoking | 10.0% (6/60) | 10.6% (7/66) | 6.5% (4/62) |
| Comorbidity | 23.3% (14/60) | 27.3% (18/66) | 51.6% (32/62) * <sup>§</sup> |
| Hypertension | 11.7% (7/60) | 15.2% (10/66) | 33.9% (21/62) * <sup>§</sup> |
| Diabetes | 3.3% (2/60) | 7.6% (5/66) | 21.0% (13/62) <sup>§</sup> |
| Initial symptoms and signs |  |  |  |
| Fever | 95.0% (57/60) | 93.9% (62/66) | 88.7% (55/62) |
| Highest temperature (°C) | 38.77 ± 0.71 | 38.66 ± 0.56 | 38.78 ± 0.75 |
| Breathlessness | 30.0% (18/60) | 37.9% (25/66) | 46.8% (29/62) |
| Chest distress or pain | 33.3% (20/60) | 28.8% (19/66) | 38.7% (24/62) |
| Fatigue or myalgia | 30.0% (18/60) | 36.4% (24/66) | 30.6% (19/62) |
| Dyspnea | 10.0% (6/60) | 12.1% (8/66) | 16.1% (10/62) |
| Respiratory failure, ARDS or sepsis | 13.3% (8/60) | 10.6% (7/66) | 17.7% (11/62) |
| Vital signs on admission |  |  |  |
| Heart rate (beats/min) | 85.5 (80.5~94) | 87 (82~98) | 89.5 (82~98) |
| Systolic pressure (mmHg) | 119 (112~130.3) | 119.5 (115~129.5) | 130 (118.3~143) <sup>§</sup> |
| Diastolic pressure (mmHg) | 76 (72.75~85) | 75 (75~83) | 77 (72~87) |
| Inflammatory and immune indexes |  |  |  |
| WBC (×10<sup>9</sup>/L) | 6.04 (4.21~9.60) | 5.77 (3.84~8.65) | 5.72 (3.73~9.25) |
| Neutrophil percentage (%) | 73.4 (61.28~84.7) | 75.2 (57.8~83.88) | 80.55 (70~89.93) * |
| Lymphocytes percentage (%) | 18.6 (9.3~31.85) | 16.5 (9.3~32.53) | 12.95 (5.68~23.2) * <sup>§</sup> |
| Monocytes percentage (%) | 5.7 (4.2~7.4) | 6.3 (4.78~8.13) | 4.75 (3.15~6.58) * |
| IL-6 (pg/mL) | 6.02 (5.21~7.82) | 6.2 (5.20~7.48) | 8.92 (6.9~13.4) * <sup>§</sup> |
| CRP (mg/L) | 22.6 (6.5~46.15) | 37.2 (11.3~80.5) | 55.3 (25.65~108.2) <sup>§</sup> |
| PCT ≥ 0.05 ng/mL | 31.7% (19/60) | 36.4% (24/66) | 64.5% (40/62) * <sup>§</sup> |
| ESR (mm/h) | 49 (39.75~65.25) | 49 (42.18~69) | 56.3 (39.5~68.15) |
| T cells count (/uL) | 737 (544~1156) | 628 (468~846) | 575 (347.8~782.8) <sup>§</sup> |
| CD4<sup>+</sup> T cells count (/uL) | 428 (291.5~750.5) | 362 (283~572) | 337.5 (166.3~452.3) <sup>§</sup> |
| CD8<sup>+</sup> T cells count (/uL) | 311 (162.5~421.5) | 236 (163~349) | 231.5 (123~290.5) <sup>§</sup> |
| Outcomes |  |  |  |
| Transfer to ICU | 15.0% (9/60) | 21.2% (14/66) | 43.5% (27/62) * <sup>§</sup> |
| Mortality | 10.0% (6/60) | 9.1% (6/66) | 50% (31/62) * <sup>§</sup> |
| Days from admission to death | 11 (4.75~11.75) | 8.5 (5.75~14.25) | 7 (4~11) |

Low hs-TnI group (n=60): hs-TnI < 2 pg/mL; Moderate hs-TnI group (n=66): 2 ≤ hs-TnI < 6.126 pg/mL; High hs-TnI group (n=62): hs-TnI ≥ 6.126 pg/mL.
<sup>#</sup> Statistically significant for Moderate hs-TnI group vs. Low hs-TnI group (P<0.05).
\* Statistically significant for High hs-TnI group vs. Moderate hs-TnI group (P<0.05).
<sup>§</sup> Statistically significant for High hs-TnI group vs. Low hs-TnI group ( $P < 0.05$ ).
Continuous data were presented by mean ± standard deviation (SD) or median (25<sup>th</sup> to 75<sup>th</sup> percentile). Categorical data were presented by percentage with number.
Abbreviations: hsTnI: High-sensitivity troponin I; ARDS: acute respiratory distress syndrome; WBC: white blood cells; IL-6: interleukin-6; CRP: C-reaction protein; PCT: procalcitonin; ESR: erythrocyte sedimentation rate; ICU: intensive care unit.

As shown in **Table 3**, increased CK-MB levels were more likely to occur in male patients and patients with current smoking. High CK-MB on admission (≥ 18 U/L) was associated with higher mortality (34.9%). High CK-MB was significantly associated with increased inflammatory levels (WBC, neutrophils, CRP and PCT) and decreased lymphocytes levels. High CK-MB was independent of acute respiratory symptoms and respiratory failure (*P*> 0.05). CK was not associated with mortality (**Supplementary table 1**).

**Table 3.** Clinical characteristics and outcomes between patients with different CK-MB levels.

| Parameters | Low CK-MB group | Moderate CK-MB group | High CK-MB group |
| --- | --- | --- | --- |
| Age (years) | 50.15 ± 13.54 | 51.19 ± 13.33 | 54.11 ± 15.77 |
| Male | 41.7% (20/48) | 67.5% (52/77) <sup>#</sup> | 74.6% (47/63) <sup>§</sup> |
| Current smoking | 0.0% (0/48) | 9.1% (7/77) | 15.9% (10/63) <sup>§</sup> |
| Comorbidity | 29.2% (14/48) | 31.2% (24/77) | 41.3% (26/63) |
| Hypertension | 12.5% (6/48) | 18.2% (14/77) | 28.6% (18/63) |
| Diabetes | 4.2% (2/48) | 10.4% (8/77) | 15.9% (10/63) |
| Initial symptoms and signs |  |  |  |
| Fever | 85.4% (41/48) | 93.5% (72/77) | 96.8% (61/63) |
| Highest temperature (°C) | 38.61 ± 0.79 | 38.71 ± 0.56 | 38.85 ± 0.71 |
| Breathlessness | 33.3% (16/48) | 32.5% (25/77) | 49.2% (31/63) |
| Chest distress or pain | 33.3% (16/48) | 29.9% (23/77) | 38.1% (24/63) |
| Fatigue or myalgia | 29.2% (14/48) | 33.8% (26/77) | 33.3% (21/63) |
| Dyspnea | 10.4% (5/48) | 13.0% (10/77) | 14.3% (9/63) |
| Respiratory failure, ARDS or sepsis | 12.5% (6/48) | 15.6% (12/77) | 12.7% (8/63) |
| Vital signs on admission |  |  |  |
| Heart rate (beats/min) | 87 (82~97) | 88 (82~98) | 86 (82~96) |
| Systolic pressure (mmHg) | 119.5 (108.5~131) | 119 (116~133) | 123 (115~140) |
| Diastolic pressure (mmHg) | 75 (72~87) | 75 (75~83.5) | 75 (75~87) |
| Inflammatory and immune indexes |  |  |  |
| WBC (×10<sup>9</sup>/L) | 5.25 (3.58~6.72) | 5.92 (3.74~8.5) | 7.83 (4.45~10.76) <sup>§</sup> |
| Neutrophil percentage (%) | 70.8 (59.03~82.6) | 74.7 (60.6~85.8) | 82 (68.7~89.6) * <sup>§</sup> |
| Lymphocytes percentage (%) | 21 (10.75~30.38) | 16.8 (9.3~32.15) | 11 (6.4~23.2) * <sup>§</sup> |
| Monocytes percentage (%) | 5.95 (4.53~7.35) | 5.7 (4.2~7.4) | 4.9 (3.2~7.4) |
| IL-6 (pg/mL) | 6.76 (5.34~10.75) | 7.36 (5.39~8.93) | 7.4 (5.5~10.42) |
| CRP (mg/L) | 26.15 (5.15~69.18) | 34.95 (10.1~76.18) | 51.7 (24.6~90.8) <sup>§</sup> |
| PCT ≥ 0.05 ng/mL | 31.3% (15/48) | 42.9% (33/77) | 55.6% (35/63) <sup>§</sup> |
| ESR (mm/h) | 52.7 (41.93~65) | 49 (39~70) | 49 (37.8~66) |
| T cells count (/uL) | 660 (547~975) | 628 (453~959) | 587.5 (366.5~846) |
| CD4<sup>+</sup> T cells count (/uL) | 390 (345~578) | 382 (270~652) | 291.5 (194~523) |
| CD8<sup>+</sup> T cells count (/uL) | 239 (171~323) | 277 (154~355) | 231 (134.5~353.5) |
| Outcomes |  |  |  |
| Transfer to ICU | 18.8% (9/48) | 18.2% (14/77) | 42.9% (27/63) * <sup>§</sup> |
| Mortality | 18.8% (9/48) | 15.6% (12/77) | 34.9% (22/63) * |
| Days from admission to death | 7 (5~15.5) | 11 (3.25~13.75) | 7 (4~8.5) |

Low CK-MB group (n=48): CK-MB < 13 U/L; Moderate CK-MB group (n=77): 13 ≤ CK-MB < 18 U/L; High CK-MB group (n=63): CK-MB ≥ 18 U/L.
<sup>#</sup> Statistically significant for Moderate CK-MB group vs. Low CK-MB group (P<0.05).
\* Statistically significant for High CK-MB group vs. Moderate CK-MB group (P<0.05).
<sup>§</sup> Statistically significant for High CK-MB group vs. Low CK-MB group ( $P < 0.05$ ).
Continuous data were presented by mean ± standard deviation (SD) or median (25<sup>th</sup> to 75<sup>th</sup> percentile). Categorical data were presented by percentage with number.
Abbreviations: CK-MB: creatine kinase-MB; ARDS: acute respiratory distress syndrome; WBC: white blood cells; IL-6: interleukin-6; CRP: C-reaction protein; PCT: procalcitonin; ESR: erythrocyte sedimentation rate; ICU: intensive care unit.

As shown in **Table 4**, increased LDH levels were more likely to occur in older patients and patients with hypertension. High LDH was significantly associated with acute respiratory symptoms and respiratory failure (*P*< 0.05). High LDH on admission (≥ 354.5 U/L) was associated with higher mortality (53.2%). High LDH was significantly associated with increased inflammatory levels (WBC, neutrophils, IL-6, CRP, PCT and ESR) and decreased immune levels (lymphocytes, monocytes, T cells and CD4^+^ T cells). Survival days were obviously shorter for deceased patients with high LDH (median: 7 days) when compared to patients with low and moderate LDH (median: 11 days, both), although it was hard to reach statistical significance because the deceased cases in the three groups were extremely uneven distribution.

**Table 4.** Clinical characteristics and outcomes between patients with different LDH levels.

| Parameters | Low LDH group | Moderate LDH group | High LDH group |
| --- | --- | --- | --- |
| Age (years) | 46.97 ± 11.2 | 50.22 ± 13.54 | 58.58 ± 15.35 * § |
| Male | 51.6% (32/62) | 67.2% (43/64) | 71.0% (44/62) |
| Current smoking | 8.1% (5/62) | 7.8% (5/64) | 11.3% (7/62) |
| Comorbidity | 27.4% (17/62) | 31.3% (20/64) | 43.5% (27/62) |
| Hypertension | 9.7% (6/62) | 18.8% (12/64) | 32.3% (20/62) § |
| Diabetes | 4.8% (3/62) | 9.4% (6/64) | 17.7% (11/62) |
| Initial symptoms and signs |  |  |  |
| Fever | 91.9% (57/62) | 92.2% (59/64) | 93.5% (58/62) |
| Highest temperature (°C) | 38.53 ± 0.69 | 38.76 ± 0.58 | 38.9 ± 0.70 |
| Breathlessness | 22.6% (14/62) | 29.7% (19/64) | 62.9% (39/62) * § |
| Chest distress or pain | 17.7% (11/62) | 28.1% (18/64) | 54.8% (34/62) * § |
| Fatigue or myalgia | 30.6% (19/62) | 34.4% (22/64) | 32.3% (20/62) |
| Dyspnea | 4.8% (3/62) | 12.5% (8/64) | 21.0% (13/62) § |
| Respiratory failure, ARDS or sepsis | 4.8% (3/62) | 12.5% (8/64) | 24.2% (15/62) § |
| Vital signs on admission |  |  |  |
| Heart rate (beats/min) | 87.5 (82~98) | 88 (82~96.75) | 87 (82~98) |
| Systolic pressure (mmHg) | 119 (115~127.3) | 119 (112.5~129) | 132.5 (119~144.3) * § |
| Diastolic pressure (mmHg) | 75 (75~85.5) | 76 (75~84) | 77 (72~87) |
| Inflammatory and immune indexes |  |  |  |
| WBC (×10<sup>9</sup>/L) | 4.81 (3.37~6.91) | 5.81 (3.59~8.27) | 8.07 (5.12~11.02) * § |
| Neutrophil percentage (%) | 63.25 (54.65~76.03) | 79.4 (68.33~83.75) # | 87.25 (74.03~91.88) * § |
| Lymphocytes percentage (%) | 27.8 (17.25~35.43) | 14.95 (10.63~23.35) # | 8.6 (5.28~17.95) * § |
| Monocytes percentage (%) | 6.85 (5.58~8.4) | 5.65 (4.65~7.1) | 3.55 (2.38~5.55) * § |
| IL-6 (pg/mL) | 5.46 (4.94~7.66) | 6.53 (5.52~7.47) | 8.63 (6.32~12.88) * § |
| CRP (mg/L) | 8.45 (2.8~28.45) | 46 (22.5~97.2) # | 64.7 (34.6~107.8) § |
| PCT ≥ 0.05 ng/mL | 17.7% (11/62) | 46.9% (30/64) # | 67.7% (42/62) § |
| ESR (mm/h) | 45 (35.85~62.5) | 48 (36.3~65) | 61 (43~70.5) § |
| T cells count (/uL) | 703 (525~1030) | 633 (467~882) | 458.5 (353.8~728.3) § |
| CD4<sup>+</sup> T cells count (/uL) | 428 (334~736) | 342 (261~501) | 287.5 (148.8~442) § |
| CD8<sup>+</sup> T cells count (/uL) | 236 (169.5~376.5) | 278 (157~368) | 184.5 (98.5~295) |
| Outcomes |  |  |  |
| Transfer to ICU | 6.5% (4/62) | 17.2% (11/64) | 56.5% (35/62) * § |
| Mortality | 3.2% (2/62) | 12.5% (8/64) | 53.2% (33/62) * § |
| Days from admission to death | 11 (11~11) | 11 (4.5~21.5) | 7 (4~10) |

Low LDH group (n=62): LDH < 253.2 U/L; Moderate LDH group (n=64): 253.2 ≤ LDH < 354.5 U/L; High LDH group (n=62): LDH ≥ 354.5 U/L.

As shown in **Table 5**, increased α-HBDH levels were more likely to occur in older patients and patients with hypertension. High α-HBDH was significantly associated with acute respiratory symptoms and respiratory failure (*P*< 0.05). High α-HBDH on admission (≥ 305 U/L) was associated with higher mortality (53.4%). High α-HBDH was significantly associated with increased inflammatory levels (WBC, neutrophils, IL-6, CRP and PCT) and decreased immune levels (lymphocytes, monocytes, T cells and CD4^+^ T cells).

**Table 5.** Clinical characteristics and outcomes between patients with different α-HBDH levels.

| Parameters | Low α-HBDH group | Moderate α-HBDH group | High α-HBDH group |
| --- | --- | --- | --- |
| Age (years) | 46.98 ± 11.28 | 50.28 ± 15.47 | 59.22 ± 13.85 * $^{\S}$ |
| Male | 54.4% (31/57) | 62.1% (36/58) | 74.1% (43/58) |
| Current smoking | 5.3% (3/57) | 10.3% (6/58) | 8.6% (5/58) |
| Comorbidity | 24.6% (14/57) | 34.5% (20/58) | 46.6% (27/58) $^{\S}$ |
| Hypertension | 7.0% (4/57) | 24.1% (14/58) $^{\#}$ | 32.8% (19/58) $^{\S}$ |
| Diabetes | 3.5% (2/57) | 10.3% (6/58) | 17.2% (10/58) |
| Initial symptoms and signs |  |  |  |
| Fever | 91.2% (52/57) | 93.1% (54/58) | 94.8% (55/58) |
| Highest temperature ($^{\circ}$C) | 38.63 ± 0.67 | 38.68 ± 0.67 | 38.93 ± 0.68 |
| Breathlessness | 26.3% (15/57) | 22.4% (13/58) | 63.8% (37/58) * $^{\S}$ |
| Chest distress or pain | 17.5% (10/57) | 27.6% (16/58) | 55.2% (32/58) * $^{\S}$ |
| Fatigue or myalgia | 21.1% (12/57) | 37.9% (22/58) | 32.8% (19/58) |
| Dyspnea | 5.3% (3/57) | 12.1% (7/58) | 22.4% (13/58) $^{\S}$ |
| Respiratory failure, ARDS or sepsis | 5.3% (3/57) | 10.3% (6/58) | 20.7% (12/58) $^{\S}$ |
| Vital signs on admission |  |  |  |
| Heart rate (beats/min) | 87 (81.5~98) | 88 (82~98) | 88 (82~98) |
| Systolic pressure (mmHg) | 119 (113.5~125.5) | 119 (115~129.5) | 131.5 (119~143) * $^{\S}$ |
| Diastolic pressure (mmHg) | 75 (73.5~84.5) | 75 (75~83) | 77 (73.5~87) |
| Inflammatory and immune indexes |  |  |  |
| WBC ($\times 10^9$/L) | 4.84 (3.42~6.27) | 5.95 (3.73~8.79) | 7.90 (4.50~10.32) $^{\S}$ |
| Neutrophil percentage (%) | 66.6 (55.85~76.05) | 78.95 (64.73~85.03) $^{\#}$ | 85.8 (70.55~90.53) * $^{\S}$ |
| Lymphocytes percentage (%) | 27.5 (15.7~35.2) | 15.05 (8.63~26.2) $^{\#}$ | 9.15 (5.55~22.35) $^{\S}$ |
| Monocytes percentage (%) | 6.8 (5.5~8.3) | 5.7 (4.68~7.75) | 3.8 (2.53~5.88) * $^{\S}$ |
| IL-6 (pg/mL) | 5.67 (5.29~7.88) | 6.06 (5.30~7.42) | 8.83 (7.32~13.44) * $^{\S}$ |
| CRP (mg/L) | 8.2 (3.05~41.35) | 33.1 (16.1~71.15) $^{\#}$ | 67.9 (37.5~112.2) * $^{\S}$ |
| PCT ≥ 0.05 ng/mL | 28.1% (16/57) | 37.9% (22/58) | 69.0% (40/58) $^{\S}$ |
| ESR (mm/h) | 44 (30.6~63.5) | 52 (41.8~71) | 57.15 (41.9~68.48) |
| T cells count (/uL) | 771 (563~1030) | 620 (385.3~823.5) $^{\#}$ | 470.5 (366.5~793.5) $^{\S}$ |
| CD4<sup>+</sup> T cells count (/uL) | 434.5 (355.3~743.8) | 328 (218~490.8) $^{\#}$ | 286.5 (164.5~487) $^{\S}$ |
| CD8<sup>+</sup> T cells count (/uL) | 255.5 (196.3~421.3) | 234.5 (154~357.3) | 230 (118.8~325.8) |
| Outcomes |  |  |  |
| Transfer to ICU | 1.8% (1/57) | 17.2% (10/58) $^{\#}$ | 48.3% (28/58) * $^{\S}$ |
| Mortality | 3.5% (2/57) | 15.5% (9/58) | 53.4% (31/58) * $^{\S}$ |
| Days from admission to death | 8.5 (6~11) | 11 (4~19) | 7 (4~10) |

Low α-HBDH group (n=57): α-HBDH < 215.8 U/L; Moderate α-HBDH group (n=58): $215.8 \leq \alpha$ -HBDH < 305 U/L; High α-HBDH group (n=58): α-HBDH ≥ 305 U/L.
$^{\#}$ Statistically significant for Moderate α-HBDH group vs. Low α-HBDH group (P<0.05).
\* Statistically significant for High α-HBDH group vs. Moderate α-HBDH group ( $P < 0.05$ ).
<sup>§</sup> Statistically significant for High α-HBDH group vs. Low α-HBDH group ( $P < 0.05$ ).
Continuous data were presented by mean ± standard deviation (SD) or median (25<sup>th</sup> to 75<sup>th</sup> percentile). Categorical data were presented by percentage with number.
Abbreviations: α-HBDH: α-hydroxybutyrate dehydrogenase; ARDS: acute respiratory distress syndrome; WBC: white blood cells; IL-6: interleukin-6; CRP: C-reaction protein; PCT: procalcitonin; ESR: erythrocyte sedimentation rate; ICU: intensive care unit.

As shown in Figure 1, the higher hs-TNI on admission was, the shorter survival days would be. hs-TNI level on admission was significantly negatively correlated with survival days (*r*= -0.42, 95% CI= -0.64∼-0.12, P=0.005). The higher LDH on admission was, the shorter survival days would be. LDH level on admission was significantly negatively correlated with survival days (*r*= -0.35, 95% CI= -0.59∼-0.05, P=0.022). CK-MB and α-HBDH were not significantly correlated with survival days (both P> 0.05).

**Figure 1.**
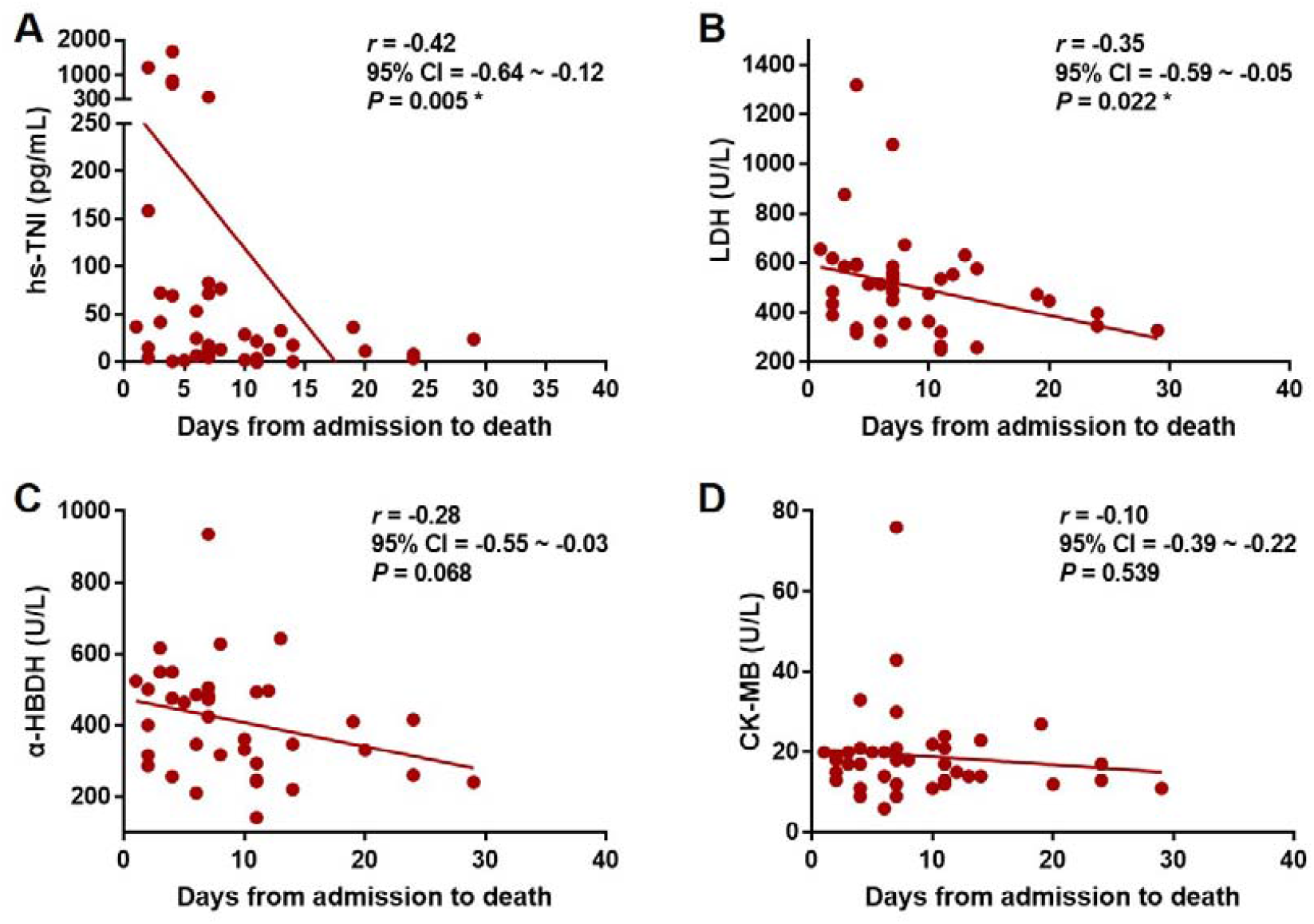
Correlations of heart injury indicators with survival days for deceased patients. A: Correlation of hs-TNI with survival days; B: Correlation of LDH with survival days; C: Correlation of α-HBDH with survival days; D: Correlation of CK-MB with survival days. * Statistically significant.

## Discussion

In this study, we first assessed the associations between heart injury indicators and mortality in COVID-19 patients. hs-TNI is a biomarker released into blood once myocardial necrosis. Aberrant hs-TNI usually specifically indicates acute myocardial infarction [8]. In our study, we found hs-TNI increase tended to occur in older patients and patients with comorbidities (especially hypertension). Approximately 11.2% COVID-19 patients happened to acute myocardial infarction on admission (aberrant hs-TNI > 28 pg/mL), which was consistent to the previous reports [9, 10]. More importantly, we found high hs-TnI on admission (≥ 6.126 pg/mL), even within the clinical upper normal limit (≤ 28 pg/mL), already can be associated with higher mortality. hs-TNI level was significantly negatively correlated with survival days. Therefore, we suggest the clinical upper normal limit of hs-TNI should be lowered for COVID-19, in order to avoid negligence of heart injury for COVID-19 patients on admission. Our suggestion also agrees with the opinion, which was issued on *The Lancet* in 2018 [11], that reclassification of hs-TNI for acute coronary syndrome in emergency departments in order to improve clinical outcomes.

CK-MB is another sensitive heart injury indicator which can also reflect myocarditis and myocardial infarction. After acute myocardial infarction, CK-MB will begin to increase at 2-4 hours, peak at 12 hours, and return to baseline within 48-72 hours [12, 13]. In our study, we found CK-MB increase tended to occur in male patients and patients with current smoking. Approximately 10.1% COVID-19 patients had CK-MB exceeding clinical upper normal limit (24 U/L) on admission. High CK-MB on admission (≥ 18 U/L), even within the clinical upper normal limit (≤ 24 U/L), already was associated higher mortality. These results proved heart injury such as viral myocarditis or myocardial infarction indeed occurred in COVID-19 on admission and predicted adverse clinical outcomes.

LDH and α-HBDH are two kinds of myocardial enzymes. LDH and α-HBDH mainly exist in myocardia and would be released into blood once myocardial injury. Although LDH and α-HBDH increases are relatively less specific in heart injury, they also play important helping roles in recognition of myocarditis or myocardial infarction [14, 15]. In our study, we found increases of LDH and α-HBDH were more likely to occur in older patients and patients with hypertension. Both high LDH and α-HBDH on admission were associated with higher mortality. In addition, we found high LDH and α-HBDH were associated with more acute respiratory symptoms such as breathlessness, chest distress or pain, dyspnea or respiratory failure on admission. This result also implied that the increases of LDH and α-HBDH might not be specific due to heart injury. LDH and α-HBDH increases might not merely be induced by heart injury, but also be induced by lung or other organs injury. Nevertheless, LDH level on admission was significantly negatively correlated with survival days.

The potential mechanism for COVID-19-caused heart injury might be virus-induced inflammatory storm. Inflammatory cells infiltration and inflammatory cytokines release can directly lead to apoptosis or necrosis of myocardial cells [16, 17]. In our study, we found high levels of heart injury indicators were associated with increased levels of inflammatory cells (WBC and neutrophils) and inflammatory cytokines (IL-6 and CRP). Therefore, we speculated that COVID-19 infection would cause heart injury such as myocarditis or myocardial infarction by inducing inflammatory cells infiltration and inflammatory cytokines storm. Early and special anti-inflammatory treatments should be undertaken by cardiologists to protect myocardium [18]. Another potential mechanism for COVID-19-caused heart injury might be angiotensin-converting enzyme 2 (ACE2)–mediated direct injury to heart. ACE2 as a human cell receptor had a strong binding affinity to the Spike protein of 2019-nCoV, namely Severe Acute Respiratory Syndrome Coronavirus-2 (SARS-CoV-2) [19]. Recent study reported ACE2 is highly expressed in heart [20]. Therefore, SARS-CoV-2 of COVID-19 might directly attack heart by binding ACE2 on myocardial cells.

Some limitations existed in this study. First, we only assessed heart injury from the perspective of serum myocardial enzymes and protein. It would be better if echocardiography and electrocardiograph were also measured in this study. Secondly, it would be better if we could assess heart injury-specific mortality in the future study with a larger sample size. Thirdly, we have not been able to obtain myocardial tissues from COVID-19 patients. Thus we currently cannot directly observe myocardial injury from the golden standard of pathology. Although the first autopsy of COVID-19 patient was completed and pulmonary pathology was obtained on Feb 16, 2020 in Wuhan Jinyintan Hospital (China), it remains a long way to remind clinicians to focus on cardiac pathology for COVID-19 infection. Our present results also can remind pathologists to pay attention to cardiac pathology in the future, especially for those deceased COVID-19 patients who had high heart injury indicators on admission.

## Conclusions

In summary, heart injury signs arise in COVID-19 on admission and would be associated with subsequently higher and earlier mortality. COVID-19 virus might attack heart via inducing inflammatory storm or attack heart directly. Clinicians must beware of heart injury indicators for COVID-19 patients, especially for those with older age, hypertension and current smoking. In order to maximally prevent or rescue heart injury-related mortality in COVID-19, cardiologists must participate in early and special managements for patients with COVID-19.

## Data Availability

The data used to support the findings of this study are available from the corresponding author upon appropriate request.

## Acknowledgements

This study was supported by The National Natural Science Foundation of China (NSFC) (81972393, 81772705, 31570775), NSFC (81630001, 81770075), Shanghai Municipal Key Clinical Specialty (shslczdzk02201) and Shanghai Top-Priority Clinical Key Disciplines Construction Project (2017ZZ02013), Sub-specialist project of Qingpu Branch of Zhongshan Hospital, Fudan university (YZK 2019-04), 2019 Hospital-level National Natural Science Foundation Incubation Project (QYP 2019-03), Science and technology development fund of Qingpu district science and technology commission in 2018 (QKY 2018-01), Academic Leader of Shanghai Qingpu District Healthcare Commission (WD2019-36), Shanghai key discipline of medicine (ZK2019B07 SZ2019-1), NSFC(81870035), Project of Shanghai municipal commission of health and family planning (201740210).

## Supplementary material

**Supplementary table 1.** Mortality between patients with different CK levels.

|  | Low CK group | Moderate CK group | High CK group | P |
| --- | --- | --- | --- | --- |
| Mortality | 17.7% (11/62) | 32.8% (21/64) | 17.7% (11/62) | 0.066 |

CK: Creatine kinase.

